# Global cognition is related to upper-extremity motor skill retention in individuals with mild-to-moderate Parkinson disease

**DOI:** 10.1101/2021.04.26.21256143

**Authors:** Jennapher Lingo VanGilder, Cielita Lopez-Lennon, Serene S. Paul, Leland E. Dibble, Kevin Duff, Sydney Y. Schaefer

**Affiliations:** School of Biological and Health Systems Engineering, Arizona State University; Department of Physical Therapy and Athletic Training, University of Utah; Discipline of Physiotherapy, Sydney School of Health Sciences, Faculty of Medicine and Health, The University of Sydney; Center for Alzheimer’s Care, Imaging and Research, University of Utah Health Sciences Center; Department of Neurology, University of Utah Hospital

## Abstract

**Background and Purpose:** Cognitive impairment has been linked to poor motor learning and rehabilitation outcomes in older adult and stroke populations, but this remains unexplored in individuals with Parkinson disease (PD). The purpose of this secondary data analysis from a recent clinical trial (NCT02600858) was to determine if global cognition was related to nine-day skill retention after upper-extremity motor training in individuals with PD.

**Methods:** Twenty-three participants with idiopathic PD completed three consecutive days of training on an upper-extremity task. For the purposes of the original clinical trial, participants trained either “on” or “off” their dopamine replacement medication. Baseline, training, and shorter-term (48-hour) retention data have been previously published. Global cognition was evaluated using the Montreal Cognitive Assessment (MoCA). Participant age, baseline performance, MoCA score, and group (medication “on”/”off”) were included in a multivariate linear regression model to predict longer-term (nine day) follow-up performance. Baseline and follow-up performance were assessed for all participants while “on” their medication.

**Results:** MoCA score was positively related to follow-up performance, such that individuals with better cognition performed better than those with poorer cognition. Participant age, baseline performance, and medication status were unrelated to follow-up performance.

**Discussion and Conclusions:** Results of this secondary analysis align with previous work that suggest cognitive impairment may interfere with motor learning in PD, and that assessing cognition could provide prognostic information about an individual’s responsiveness to motor rehabilitation for a number of clinical populations.

## INTRODUCTION

Despite clear evidence that upper extremity motor deficits in Parkinson disease (PD) negatively impact activities of daily living,^1^ most rehabilitation research and clinical practice for PD focuses on gait and balance problems. When prescribed, however, motor rehabilitation can improve upper extremity movement patterns and physical function,^2^ although some patients show marked gains following therapeutic intervention, while others do not (e.g.,^2^). The ability to predict therapeutic responsiveness could help therapists streamline and personalize treatments. However, most predictive tools or models of post-intervention motor outcomes are time- and cost-intensive (e.g., annual clinical measures, neuroimaging, etc.).

In contrast, cognitive assessment may be a feasible, brief, and relatively inexpensive tool for gaining insight to an individual’s motor learning capacity (see^3^). For example, global cognitive status has been shown to predict gains in walking speed following standard-of-care physical therapy independent of primary diagnosis,^4^ but the relationship between global cognitive measures and clinical upper-extremity outcomes in PD has not been explored. In a recent randomized clinical trial in individuals with mild-to-moderate PD (clinicaltrial.gov registration number NCT02600858),^5^ motor practice while “on” dopamine replacement medication (i.e., levodopa) improved 48-hour retention of a functional upper extremity motor task compared to practice “off” dopamine replacement medication. The purpose of the present study was to perform secondary analyses of these data to evaluate whether cognition was related to skill learning in the upper extremity.

## METHODS

### Participants

Twenty-three adults aged >50 years old with a confirmed diagnosis of PD were included in this secondary analysis of data from a previously published randomized clinical trial (clinicaltrials.gov registration number NCT02600858).^5^ Exclusion criteria included prior surgical treatment of PD (e.g., deep brain stimulation), dementia (Montreal Cognitive Assessment (MoCA) < 18), and the presence of concomitant neurological conditions. Included participants must have been taking dopamine replacement medications. The clinical trial protocol required half the participants (n=12) to complete upper extremity motor training while continuing to take their prescribed dose of levodopa medication; the other half (n=11) skipped their first dose of medication each day of motor training such that they were “off” medication following overnight withdrawal. These participants took their remaining daily doses after they completed the motor training each morning. Details regarding dopamine medication and other participant characteristics have been previously reported.^5^

Global cognition was measured using the MoCA, a brief cognitive screening tool in which scores range from zero to 30; a score of 26 (or higher) is considered normal. To evaluate upper extremity dexterity, participants completed the Nine-hole peg test (a timed clinical measure of dexterity in which faster trial times indicate better performance) and a timed experimental task that simulates buttoning a shirt unimanually (i.e., dexterity task). Participants also completed the motor subsection of the Movement Disorder Society-Unified Parkinson Disease Rating Scale (MDS-UPDRS) to evaluate the severity of PD motor signs; higher scores indicate greater severity. To evaluate depressive symptoms, participants completed the Geriatric Depression Scale, a self-report rating tool consisting of 15 items and a score of four or lower is considered normal. Participants self-reported hand dominance. All participant characteristic data were collected while the participants were “on” their prescribed dose of dopamine replacement medication, regardless of which group they were randomized to for training (“on” vs. “off” medication).

### Upper extremity motor training

As described previously,^5^ the motor training protocol required participants to complete a familiarization trial, then 50 training trials each day for three consecutive days. Participants were then re-tested two and nine days later; the two-day follow-up was the stated primary outcome of the clinical trial, thus, only the longer-term nine-day follow-up was included in this analysis.

The motor task used in this study was designed to mimic an activity of daily living and has been validated against subjective and objective measures of daily functioning in other cognitively impaired samples^6^; thus, it represents a task that would be practiced in clinical motor rehabilitation. The experimental apparatus comprised three ‘target’ cups placed 16 cm from a center ‘home’ cup at 45, 90, and 135 degrees. Participants were asked to use a plastic spoon held in their nondominant hand to collect two raw kidney beans from the home cup and transport them to one of the three target cups. The nondominant hand was used to ensure the task was not overlearned and to avoid potential confounds of a ceiling effect. Participants were instructed to move first to the target cup ipsilateral to the nondominant hand, then to the middle cup, then to the contralateral cup, repeating this pattern four more times. Thus, each trial consisted of 15 reaches. The primary measure of performance was *trial time*, which began when the participant picked up the spoon and ended when they completed all reaching movements and placed the spoon back onto the table; thus, lower trial times indicated better performance. Dropping beans, transporting an incorrect amount, or moving to the wrong target were counted as errors, and the participant could not continue until the error was corrected; thus, errors were reflected in longer trial times. Participants were not provided with performance feedback but could explore different movement strategies to optimize performance (i.e., discovery learning). As noted previously, each training session consisted of 50 trials (i.e., 750 reaches per session), and participants completed three training sessions over three days (1/day), totaling 2,250 reaches. This dose of training on this task was selected based on previous feasibility and efficacy studies in other clinical populations, which are referred to in (Paul et al., 2020).

### Statistical analysis

JMP Pro 14.0 (SAS Institute Inc., Cary NC) was used for all statistical analyses. To examine whether global cognition was related to the amount of learned motor skill, MoCA scores were included in a multivariate linear regression model as a predictor of nine-day follow-up performance (α=0.05), along with baseline motor performance, performance on the motor subsection of the MDS-UPDRS, and age as covariates. The motor portion of the MDS-UPDRS was included to control for the severity of PD motor signs; baseline motor performance was measured as the first trial of the first motor training session. Although we did not have a specific hypothesis regarding the effect of dopamine replacement medication on learning, we also included the variable of group (“on” vs. “off” medication) as a covariate to control for any confounds of dopamine replacement medication status on the primary outcome.

## RESULTS

As shown in Table 1, the average MoCA score was >26, suggesting that the sample overall was predominantly cognitively intact; however, these scores ranged between 23 to 30, indicating variability within the sample. Participants demonstrated mild PD symptoms and disease severity (median Hoehn and Yahr stage was 2.0), consistent with their objective motor assessment data. Nine-hole peg test scores were consistent with previously reported values in PD^7^, and dexterity task scores were two times worse than those of healthy older adults from previously reported data.^8^

**Table 1.** Sample (n=23) characteristics.

| Characteristic | Mean (±SD) | Range (Minimum-Maximum) | Std. Error | 95% Confidence interval |
| --- | --- | --- | --- | --- |
| **Age (years)** | 71.07 (±6.88) | 50.5-80.3 | 1.43 | 68.09-74.05 |
| **Education (years)** | 16.30 (±2.51) | 12-20 | 0.52 | 15.22-17.39 |
| **GDS (0-10)** | 2.76 (±3.1) | 0-10 | 0.55 | 0.09-0.39 |
| **MDS-UPDRS 3 (0-132)** | 30.53 (±8.84) | 13-55 | 1.8 | 26.79-34.34 |
| **MoCA (0-30)** | 26.35 (±2.04) | 23-30 | 0.40 | 25.61-27.25 |
| **9 HPT-ND (seconds)** | 26.01 (±3.97) | 3.97 | 0.83 | 24.29-27.72 |
| **UE Dexterity Task (seconds)** | 102.81 (±54.37) | 54.37 | 11.33 | 79.30-126.32 |
| **Motor task Baseline (seconds)** | 64.1 (±9.4) | 46.72-84.91 | 1.97 | 60.24-67.96 |
| **Motor task 9-day Follow-up (seconds)** | 55.0 (±11.8) | 36.38-77.62 | 2.46 | 50.18-59.82 |
| **Abbreviations:** |  |  |  |  |
| SD = Standard deviation |  |  |  |  |
| GDS = Geriatric Depression Scale |  |  |  |  |
| H&Y = Hoehn and Yahr |  |  |  |  |
| MoCA = Montreal Cognitive Assessment |  |  |  |  |
| MDS-UPDRS 3 = Movement Disorder Society – Unified Parkinson Disease Rating Scale Motor Portion |  |  |  |  |
| 9 HPT-ND = Nine Hole Peg Test Non-Dominant Hand (prior to intervention) |  |  |  |  |
| UE Dexterity Task = Upper Extremity simulated buttoning dexterity task (nondominant hand) |  |  |  |  |

Linear regression model results indicated that MoCA score predicted nine-day follow-up performance (β=-2.91; 95% CI [−5.05, −0.77], p=0.0105), whereas participant age (β=-0.18; 95% CI [−0.88, 0.47], p=0.52), severity of PD motor signs (β=0.14; 95% CI [−0.41 0.69], p=0.59), and medication status group (β=2.33; 95% CI [−2.19, 6.86], p=0.29) did not (see Table 2).

**Table 2.** Multivariate regression results.

| **Name** | **Estimate ($\beta$)** | **Std. Error** | **t-value** | **p-value** |
| --- | --- | --- | --- | --- |
| **Intercept** | 113.12 | 40.48 | 2.79 | 0.0125* |
| **Group** | 2.33 | 2.14 | 1.09 | 0.2923 |
| **Age** | -0.18 | 0.33 | -0.54 | 0.5949 |
| **Baseline motor task performance** | 0.44 | 0.26 | 1.70 | 0.1081 |
| **MDS-UPDRS 3** | 0.14 | 0.26 | 0.54 | 0.5988 |
| **MoCA** | -2.91 | 1.01 | -2.88 | 0.0105* |

*Notes:*
*Based on 23 Observations*
*Degrees of freedom (5,17).*
**MDS-UPDRS 3 = Movement Disorder Society – Unified Parkinson Disease Rating Scale Motor Portion (On medication and Off medication)**

## DISCUSSION

The purpose of this secondary analysis was to determine whether cognition was related to upper extremity motor skill learning in individuals with PD. Results indicated that MoCA score predicted follow-up performance of a functional upper extremity motor task nine days after the last practice session, more so than baseline performance and age (regardless of “on”/”off” medication training groups). These findings align with previous work that suggest cognitive testing can be used to predict rehabilitative outcomes^9^ and support that global cognition may be a useful tool to predict motor learning in clinical populations.^10^ Although the MoCA was used to evaluate global cognition in this study, there is existing evidence that supports the value of assessing specific cognitive domains as predictors of motor learning.^9^ We further acknowledge that this study focused on upper extremity motor skill specifically, but future work should investigate which (and to what extent) specific cognitive impairments (e.g., visuospatial, delayed memory) interfere with learning different types of skills.

In the published clinical trial,^5^ there was a modest effect of medication status during training (i.e., “on”/”off” medication while practicing the task) between baseline and 48-hour follow-up task performance, such that the “on” medication group performed significantly better at this short-term retention period than did the “off” medication group. These results were interpreted to indicate that being “on” dopamine replacement medications may facilitate short term retention of motor skill. However, this secondary analysis indicates that the group difference was no longer present by the ninth day of retention, likely due to the modest effect of medication status on training previously observed. Instead, global cognition (which was not originally considered in the parent clinical trial) was a significant predictor of motor task performance well after training had been completed (nine days later), regardless of whether training had occurred “on” or “off” dopamine replacement medication. Indeed, dopamine replacement may be insufficient to offset the breadth of cognitive deficits associated with PD,^11^ and the short duration in which participants in the “off” group were withdrawn from their medication for training (relative to the nine-day duration of retention) may explain the lack of effect of group in this secondary analysis.

One limitation of this study is that the participants in the “off” medication group resumed their regularly prescribed dopamine replacement therapy after training each day and throughout the nine-day retention period; thus, we are unable to discern potential effects of medication adherence or withdrawal on motor skill consolidation and retention, which are also critical periods for motor learning, as well as acquisition. We also acknowledge that this study was not designed to directly test if global cognition would be predictive of clinical rehabilitation motor outcomes in individuals with PD, since it only evaluated the amount of skill retained over a period of nine days. Performance of the functional upper extremity task used in this study has, however, been associated with subjective and objective measures of daily functioning in individuals diagnosed with Mild Cognitive Impairment.^6^ Another limitation to this study is inherent to the MoCA itself, as it is a very gross screening tool that does not thoroughly assess the function of each cognitive domain nor is it validated to measure the function of individual cognitive domains. Nonetheless, the MoCA has been shown to have reasonable accuracy for screening neurocognitive disorders,^12^ suggesting that it may be valuable for clinical motor rehabilitation as well. Lastly, participants in this study trained on the motor task with the non-dominant upper extremity to ensure task novelty and avoid a ceiling effect. Future work should consider training of the more affected upper extremity and test the clinical utility of a more comprehensive cognitive battery in predicting motor rehabilitation responsiveness in individuals with PD.

## CONCLUSIONS

Results of this study suggest that global cognition may influence upper-extremity rehabilitation for individuals with PD. These findings align with previous work that suggest cognitive testing can be used to predict rehabilitative outcomes and support that global cognition may be a useful tool to predict motor learning in clinical populations. Future work should investigate how impairments in specific cognitive domains affect motor learning and rehabilitation outcomes in individuals with PD and other neurologic populations.

## Data Availability

All data will be made available upon reasonable request.

